# A Tri-light Warning System for Hospitalized COVID-19 Patients: Credibility-based Risk Stratification under Data Shift

**DOI:** 10.1101/2022.12.11.22283309

**Authors:** Chuanjun Xu, Qinmei Xu, Li Liu, Mu Zhou, Zijian Xing, Zhen Zhou, Changsheng Zhou, Xiao Li, Rong Wang, Yanjun Wu, Jiangtao Wang, Longjiang Zhang, Xianghao Zhan, Olivier Gevaert, Guangming Lu

**Author notes:** Contributing authors. These co-corresponding authors contributed equally to this work (Chuanjun Xu, Qinmei Xu, Li Liu, Xianghao Zhan, Olivier Gevaert, Guangming Lu).

## Abstract

**OBJECTIVE:** To develop a tri-light warning system for the early warning of novel coronavirus pneumonia (COVID-19) and stratification of patients.

**MATERIALS AND METHODS:** The system extracts radiomic features from CT images and integrates clinical record information to output a prediction probability and credibility of each prediction. It classifies patients in the general ward into red (high risk), yellow (uncertain risk), and green (low risk) labels. The system was tested using a multi-center cohort of 8,721 patients.

**RESULTS:** The system demonstrated reliability and performance validation under data distribution shifts, and was applicable to both the original strain and variant strains of COVID-19.

**DISCUSSION:** The tri-light warning system has the potential to improve patient stratification performance and identify epidemiological risks early, thus allowing for timely treatment and optimization of medical resource allocation.

**CONCLUSION:** The tri-light warning system based on conformal prediction is a reliable and effective method for the early warning and stratification of COVID-19 patients.

## 1 Introduction

Coronavirus disease 2019 (COVID-19) continues to spread and has caused over 526 million confirmed cases and over six million deaths by 29 May 2022 [1]. Various mutant strains emerged with increased infectiousness (e.g. Omicron) or morbidity (e.g. Delta) when compared with the previously observed strain in the pandemic [2] [3]. Near-capacity hospital and intensive care unit use were commonly reported during the peaks of pandemic waves [4]. Therefore, reliable, generalizable and sustainable methods for timely identification of high-risk patients are crucial for clinical decision making and efficient allocation of resources in the context of existing and emerging virus strains[5].

COVID-19 is primarily characterized by pulmonary inflammatory lesions, where computed tomography (CT) feature assessment by radiologists is used for treatment evaluation [6] [7] [8] [9]. However, current processes are often subjective and unable to accurately predict the disease progression, leading to significantly increase the workload for radiologists. Therefore, an automated and quantitative analytical method of analyzing CT images is urgently needed to provide more objective and reliable evaluation for better determinination of disease progression.

Recent studies have confirmed that an image-based AI prognostic prediction model can play a supporting role in determining disease progression of COVID-19 patients [10][11][12][13]. Clinical information such as sex, age, symptoms, comorbidities and laboratory values of patients are expected to be used for more accurate assessment of prognosis [14][11][15]. For instance, AI-based models utilizing CT radiomic features and clinical indicators can predict high-risk events during hospitalization and events such as admission length, which demonstrated the value of imaging and clinical features in prognostic prediction [16][17][18]. However, daunting challenges remain in the early warning of disease progression and its decision making with confidence for COVID-19 cases.

Current research efforts do not address the generalizability of models under data distributions caused by mutant strains and variability related to the hospital setting [19][20]. Because of the changing status of virus mutation, models developed based on existing strains may no not be applicable to new strains. In addition, unlike evidence gained from small cohorts[21][22], we emphasize the necessity of multi-center evaluation for testing model generalization. Clinical centers are often equipped with various medical devices, protocols and resources. For instance, the field hospitals and permanent hospitals for COVID-19 patients can have different conditions of equipment, medical personnel, and protocols. Such difference caused by the hospital types increases the difficulty to measure model performance. Further, existing studies lack the analysis of prediction reliability, i.e., how much confidence we have for a particular prediction and is high prediction confidence related to high prediction accuracy [23][24][25].

To address these challenges, we collected PCR-confirmed COVID-19 patients from 40 hospitals across China, developing an end-to-end flexible tri-light warning system to predict the potential requirement of ICU care of patients (Figure 1). This system is based on conformal prediction that calculates the credibility of each prediction. Briefly, this system stratifies patients into three categories: a. Red: high probability with high credibility, represents high risk patients that need ICU care within 28 days; b. Green: low probability with high credibility, represents low risk patients; c. Yellow: high/low probability with low credibility, represents patients with uncertainty of risk that need further monitoring.

**Fig. 1.**
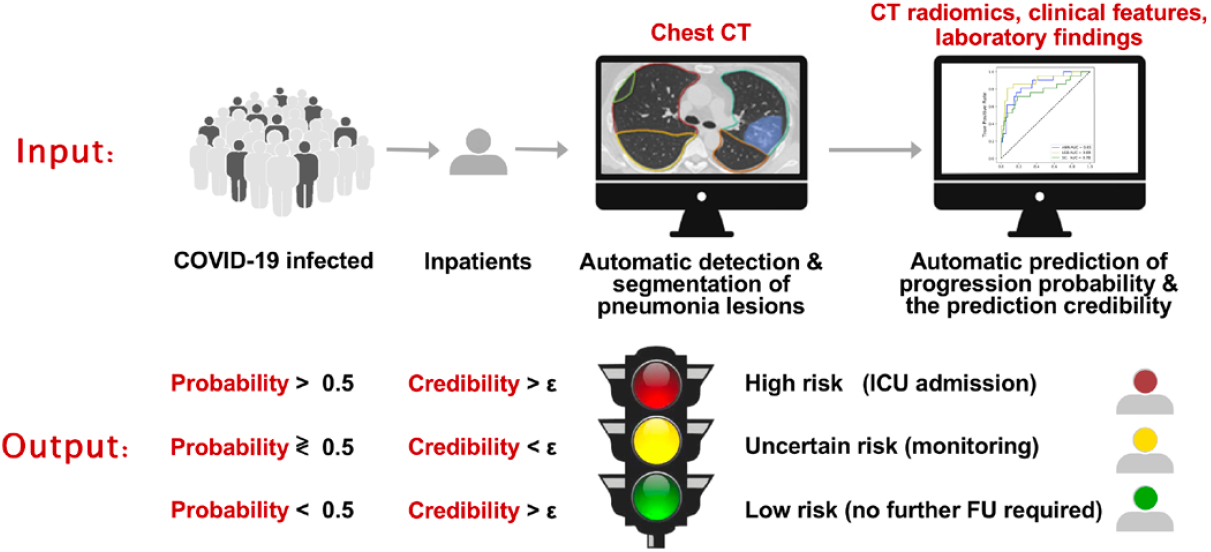
Workflow of the proposed Tri-light Warning System for hospitalized COVID-19 patients. First, the system automatically detects and segments the pneumonia lesions on the input chest CT images of patients at admission, and extracts the radiomic features from the lesion. Second, the radiomic features, clinical features, and laboratory findings of patients were combined to automatically predict whether the patient’s condition will progress and require ICU care within 28 days, and the prediction credibility was also given. Finally, the system classifies patients into high-risk, low-risk and uncertain risks according to the progression probability and the prediction credibility, aiming to provide closer medical monitoring for high-risk and uncertain patients, and dispatch medical resources in advance to ensure timely treatment of high-risk patients. Particularly, medical institutions can adjust the credibility threshold *ϵ* of the system according to the virus characteristics (mortality, infectivity, etc.) and local medical resources, so as to flexibly change the proportion of high-risk patients output by the system and the reliability requirement of prediction. CT = computed tomography; ICU = intensive care unit; FU = follow-up

The major contributions of this study can be summarized as follows: first, based on conformal prediction, we developed an early warning system for hospitalized COVID-19 patients. This system quantifies the credibility of each prediction when calculating the probability of progression, providing important clinical guidance for patient management and treatment mode selection. Second, we proposed a flexible tri-light warning strategy, which can change the proportion of high-risk patients from model output according to local medical resource allocation and virulence of virus, helping to optimize the allocation of medical resources, thus achieving closer monitoring and timely treatment for high-risk patients. Third, we collect a multi-center cohort from 40 hospitals in China (n=8721) for systematic evaluation. To assess the mutational effect on virus, we perform analysis based on COVID-19 patients infected with the original strain, and validate the performance on data from other hospitals with different strains (i.e. delta and omicron, n=270) to evaluate the generalization of the model.

The remaining sections of this paper are organized as follows. Section II discusses related work. The architecture and implementation detail of the proposed method are presented in Section III, followed by the experimental results in Section IV. Finally, the discussion and concluding remarks are given in Section V.

## 2 BACKGROUND: AI-enabled COVID-19 studies

Predicting patient outcomes with COVID-19 at an early stage is crucial to optimize the clinical care and medical resource management [26]. Multiple AI models based on machine learning (ML) and deep learning have been proposed to address this task. Example models estimated mortality risk in patients with suspected or confirmed COVID-19 [19][20][22][24][25]. Other models aimed to predict progression to a severe or critical state [21][23].There are also efforts to predict the length of hospital stay [27][15]. The most common prognostic predictors included age [19][20][28][23][29][15][30], sex [31][20][29][30], comorbidity (including hypertension, diabetes, cardiovascular disease, respiratory disease) [20], lymphocyte count [19][22], and also radiomic features derived from CT images [21][24][15][32],.

Recently developed models share a similar perspective with our research and show the potential value of clinical application. A study proposed a clinical risk score to predict the occurrence of critical illness in hospitalized patients with COVID-19 from clinical and radiology report data (AUROC: 0.88) [33]. Another study developed a recurrent neural network-based model to predict the outcomes of patients with COVID-19 by using available electronic health records on admission to hospital (AUROC: 0.85-0.93) [4]. Also, our previous study used 3,522 PCR-confirmed COVID-19 inpatients from 39 hospitals and performed CT-based analysis combined with electronic health records and clinical laboratory results with prognostic estimation for the rapid risk stratification (AUROC 0.916-0.919) [16].

However, in AI-based applications, the reliability of predictions is significant for assisted decision and risk control in real-world applications: unreliable prediction can interfere with the clinicians’ decision and may lead to misdiagnosis put on huge pressure on the patient families. Therefore, knowing how much confidence is associated with a prediction made by the model is important for the decision making process for the clinicians [34]. PROBAST analysis indicates that the majority of proposed models above are at a high risk of bias, and their reported performance is probably optimistic [26][35]. Unreliable predictions could cause more harm than benefit in guiding clinical decisions. Therefore, current AI-enabled models have not been validated or implemented outside of their original study sites, which are therefore not recommended in clinical practice [26][4]. To address this challenge, we developed prognostic prediction models based on a large, heterogeneous, real-world data set in a newly proposed conformal prediction framework [36, 37]. The models not only provide high prediction performance in predicting whether a patient in the general ward will be admitted to the intensive care units (ICUs) under distribution drifts, but also provide the users with prediction reliability information. Based on the prediction reliability information given by the conformal prediction framework, a tri-light warning system is introduced to enable the users to adapt health policies for ICU resource allocation 1. This framework also has potential applications in future outbreaks or other pandemics.

## 3 Methods

### 3.1 Patient cohort

The data in this study were collected from 40 hospitals in China (n=8721). Patients selection followed the inclusion criteria: (a) RT-PCR confirmed positive severe acute respiratory syndrome coronavirus (SARS-CoV-2) nucleic acid test; (b) baseline chest CT examinations and laboratory tests on admission; (c) short-term prognosis information (discharge or admission to ICU). Along with the exclusion criteria, we collected 3646 patients for analysis, including four cohorts. First, a training cohort (n = 1451) is set for model development, which included patients from 17 hospitals. Second, we performed model parameter tuning on a validation set (n = 662) which consisted of patients from nine independent medical centers. Third, we assessed the performance of models on an external test set (n = 1263) with higher rate of ICU admission from Huoshenshan (HSS) field Hospital, an emergency specialty field hospital designed to treat people with COVID-19 in Wuhan, Hubei, China. In addition, we built a specific test set (n = 270) based on COVID-19 Delta and Omicron variants to evaluate the generalization of models. An overview of the patient cohorts is summarized in Fig. 2.

**Fig. 2.**
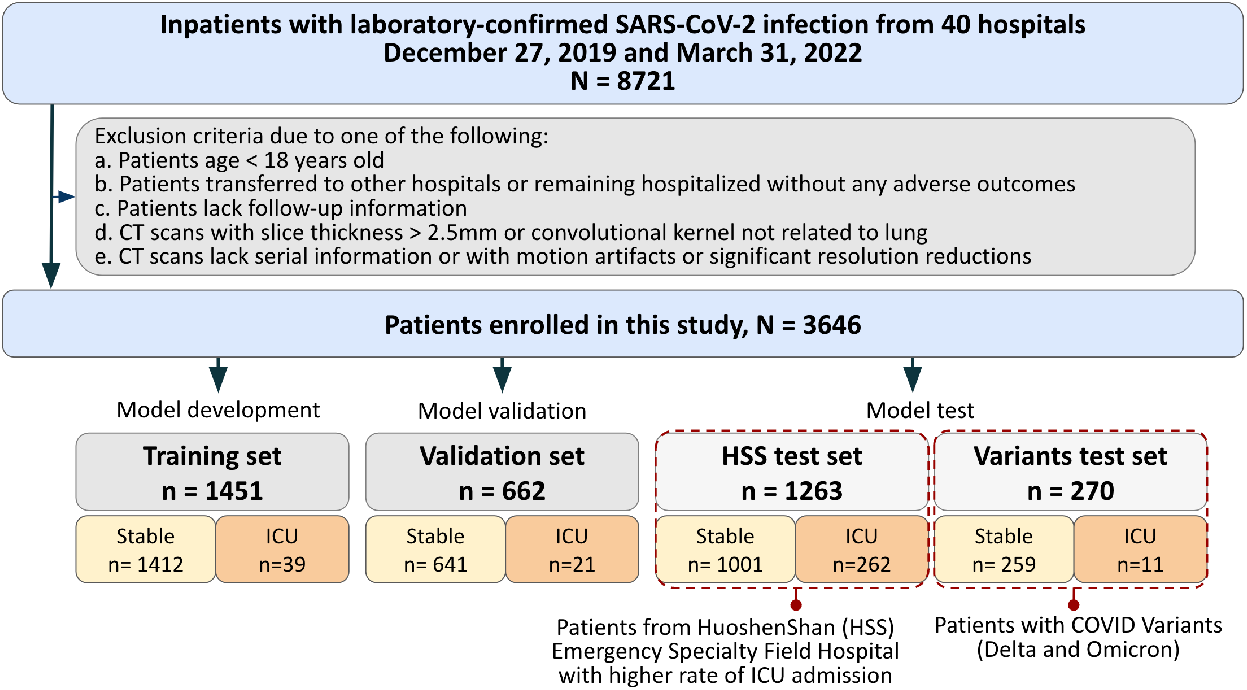
The summary of the patient cohorts and exclusion criteria. The models were trained on the training set and the model hyperparameters were tuned on the validation set. Upon hyperparameter tuning and optimizing the feature selection approach on the validation set, the training set and validation set are combined to train the ultimate models. The ultimate models are tested on the two test cohorts under data distribution drifts: the Huoshenshan field hospital test set and the Delta and Omicron variant data set.

### 3.2 CT protocols

Patients took baseline CT scans within three days after admission [38]. Chest CT scans were performed using ≥ 16 slice multidetector CT scanners (Aquilion ONE / Aquilion PRIME / BrightSpeed / BrightSpeed S / Brilliance 16 / Brilliance 64 / Discovery CT750 HD / eCT / Fluorospot Compact FD / HiSpeed Dual / iCT 256 / Ingenuity CT / Ingenuity Flex / LightSpeed VCT / LightSpeed 16 / NeuViz 16 Classic / Optima CT520 Series / Optima CT540 / Optima CT680 Series / ScintCare CT 16E / Sensation 64 / SOMATOM Definition AS+ / SOMATOM Definition Flash / uCT 510) without use of iodinated contrast agents. To minimize motion artifacts, patients were asked to hold their breath, then axial CT images were acquired during end-inspiration. The CT scan protocols were as follows: tube voltage, 100 − 120 kVp; effective tube current, 110 − 250 mAs; detector collimation, 16 − 320 × 0.625 − 2.5 mm; slice thickness, 0.625 − 2.5 mm; pitch, 0.8 − 1.375. Based on the raw data, the CT images were reconstructed by iterative reconstruction technique if possible.

### 3.3 Data collection and preprocessing

Our multi-modal data for each patient included: 1) Clinical data based on electronic Health Record (EHR): (a) demographics: age and gender; (b) comorbidities: coronary heart disease, diabetes, hypertension, chronic obstructive lung disease (COPD), chronic liver disease, chronic kidney disease, and carcinoma; and (c) clinical symptoms: fever, cough, myalgia, fatigue, headache, nausea or vomiting, diarrhea, abdominal pain, and dyspnea on admission. To extract the clinical data from the free-text EHR in Chinese, we developed a rule-based language processing algorithm. Firstly, the clinical descriptions are segmented from EHR by splitting the paragraphs with subtitles. Then, we established a keyword list containing all the descriptions associated with a particular clinical feature, such as ‘fever’,’cough’. Simply using a regular expression to match keywords is not practical. An example of the clinical description is: “this patient had fever and cough three days ago, and he had no diarrhea or vomiting, and today he is transmitted to this hospital without fever”. Since the symptoms can progress/recover, the same keywords have different meanings with negation and thought groups. Therefore, we designed a voting rule to extract the clinical data: by breaking down the clinical descriptions with commas, the keywords can be matched into several thought groups. If a thought group begins with negation, the keywords appeared vote zero to their encoding values; Otherwise, the keywords appeared vote one. The votes are summed up and compared with zero, and then the sum will convert into a boolean value and be viewed as the encoded value for a specific clinical feature.

2) Laboratory test: (a) blood routine: white blood cell (WBC) count (× 10^9^*/L*), neutrophil count (× 10^9^*/L*), lymphocyte count (× 10^9^*/L*), platelet count (× 10^9^*/L*), and hemoglobin (*g/L*); (b) coagulation function: prothrombin time (PT) (*s*), activated partial thromboplastin time (aPTT) (*s*), and≤ D-dimer (*mg/L*); (c) blood biochemistry: albumin (*g/L*), alanine aminotransferase (ALT) (*U/L*), aspartate Aminotransferase (AST) (*U/L*), total bilirubin (*mmol/L*), serum potassium (*mmol/L*), sodium (*mmol/L*), creatinine (*µmol/L*), creatine kinase (CK) (*U/L*), lactate dehydrogenase (LDH) (*U/L*), *α*-Hydroxybutyrate dehydrogenase (HBDH) (*U/L*); (d) infection-related biomarkers: C-reactive protein (CRP) (*mg/L*). Patients took laboratory test on the date of admission in training set, validation set, and the Variants test set, while patients in HSS test set receiving laboratory test within three days after admission due to the centralized outbreak in Wuhan and the limited medical resources. To alleviate missing values that occurred in records, we applied median imputation on the lab data when a missing rate was ≤50%, which has been validated effective in the previous study [16]. Each inpatient received laboratory tests within 48h after admission and only clinical data on or prior to the date of the CT were used for prediction.

3) CT radiomics: a commercial deep-learning AI system (Beijing Deepwise & League of PhD Technology Co. Ltd) was first used to detect and segment the pneumonia lesion, and two radiologists (Q.M.X. and C.S.Z.) checked the results of the automatic segmentation. Then, pyradiomics (v3.0) running in the Linux platform was adopted to extract radiomic features (1652 features per lesion). Next, for a given patient and for each radiomic feature, we summarized the distribution of the feature values across all the lesions for the patient by several summary statistics (mean, median, standard deviation, skewness, the first quartile, the third quartile) and the number of lesions. Finally, a total of 9913 quantitative radiomic features were extracted from CT images for each patient.

### 3.4 Feature engineering

To address the imbalance in the data set and high dimensionality of the feature space before modeling, several different feature engineering approaches were applied to select/weigh the features and augment the minority cases:

1. synthetic minority oversampling technique (SMOTE): SMOTE over-samples the minority class by synthesizing new minority data. Under the assumption that data close in the feature space are similar in their labels, SMOTE randomly selects a pair of minority-class data, draws a line between them in the feature space and finds a random point along the line segment as the new synthetic minority-class data. SMOTE was implemented with the Python package imblearn (version: 0.6.2);
2. feature selection based on shrunken centroids (SC): SC is an algorithm derived from the nearest centroids (NC) [39]. However, SC further attenuates the noisy features which do not have much class-related information. The SC feature selection works with the following steps (assume the original feature space has a dimensionality of *D*): Firstly, in the original feature space, the centroids 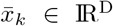 of each class (1, 2, …, *K*) and the overall centroid *µ* of all samples are computed (*Z*:((*x*_1_, *y*_1_), …, (*x*_*n*_, *y*_*n*_))). Here *C*_*k*_ denotes the sample set with label *k*, and *n*_*k*_ denotes the number of samples with label *k*.

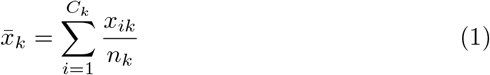

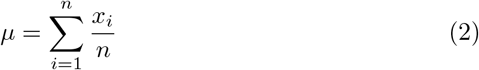 Secondly, the pooled within-class standard deviation is calculated as an unbiased estimator of the standard deviation of the overall distribution, and then the contrasts between class centroids and the overall centroid are standardized by the pooled within-class standard deviation:

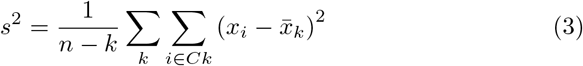

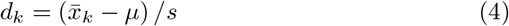 Finally, the contrasts are shrunken with a threshold denoted as ∆, which is a hyperparameter:

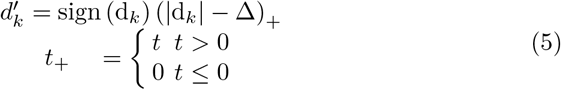 The feature selection effect is exerted by the threshold ∆: if *d*_*jk*_, the contrast of a *j*-th feature of class *k*, has an absolute value which is smaller than the threshold ∆, the corresponding feature will be considered not informative enough for classification. Therefore, the contrast in this feature will be shrunken to zero and the noisy feature is therefore removed, which also lowers the data dimensionality.
3. feature selection based on Lasso (Lasso): a logistic regression model is fitted with L1 penalty. By adding different strengths of L1 penalty, different numbers of features will be given zero coefficients in the logistic regression and the features with non-zero coefficients will be selected to reduce the dimensionality of the feature space;
4. feature weighing based on principal component analysis (PCA): PCA performs the covariance analysis and finds the principal components which maximize the variance of the data projections. By projecting the features onto the principal components, the dimensionality of the original feature space is reduced and the information is compressed in the projections on the principal components. It should be noted that generally, PCA only weighs the features but does not mask features or perform feature selection. Lasso and PCA were implemented with the scikit-learn package (version: 0.21.3).

In this study, SMOTE was first implemented to cope with the class imbalance and then one of the feature selection/weighing methods (SC/Lasso/PCA) was used. The hyperparameters associated with these algorithms that have been tuned in this study include: the threshold ∆ for SC feature selection, the strength of the L1 penalty for Lasso feature selection *C*, and the number of principal components selected for PCA.

### 3.5 Prediction model development and evaluation

In this study, the task is to predict whether a patient admitted in the general ward will be admitted to the intensive care unit (ICU) within 28 days. To address this classification task, we concatenated the CT radiomic features, clinical features (including the demographics, clinical symptoms and commorbidities) and the lab test features. Then, we leveraged the shrunken centroids (SC)[36, 39], Light Gradient Boosting Machine (LGB)[40] and artificial neural network (ANN) [41]. These algorithms are chosen because they are the representatives of different classification rationales: SC classifies samples based on the the similarity in the Euclidean feature space (as a modified version of the nearest centroids algorithm) [36, 39], LGB is a tree-based ensemble-learning algorithm with the boosting ensemble-learning strategy [40], and ANN is a representative of the deep learning technology based on the gradient descent in minimizing the binary cross-entropy loss function [41]. The SC was implemented with Python 3.7 [36]. The LGB was implemented with the Python package lightgbm. The ANN was implemented with the Python package scikit-learn (version: 0.21.3).

The hyperparameters are tuned based on the performance on the validation set in this study and the types of hyperparameters tuned are listed as follows: 1) for SC: the threshold ∆; 2) for LGB: the learning rate, the maximum depth of trees and the number of leaves of the trees; 3) for ANN: the number of hidden layers and the numbers of hidden units for each hidden layer. It should be mentioned that the classifier hyperparameters are tuned in combination with the feature selection methods and feature selection hyperparameters on the validation data set.

### 3.6 Analysis of important features in decision

To provide users with more explainable and interpretable decision making based on the model, we investigated the important features which support the decision-making process. The importance is measured by the absolute value of the shrunken contrasts 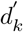. After the soft-thresholding operation, the irrelevant and noisy features will be set to zero, and the other features will be reduced by the threshold ∆. Therefore, the valuable features will remain high in the vector 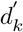, and their absolute values directly indicate the weights in the prediction process. To ensure the robust important features are investigated, with 30 times of bootstrapping, we calculated the mean absolute values of shrunken contrast for each feature on the shrunken centroid for class ‘positive’. The higher the mean absolute value of the shrunken contrasts indicates that the corresponding feature is given more weight by the model. Meanwhile, the signs of the shrunken contrast for the features indicate the positive/negative association with a positive prediction: i.e., for the *j*-th feature, if 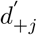 is positive, a higher feature value contributes to a positive prediction, while if 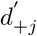 is negative, a lower feature value contributes to a positive prediction. Additionally, we also investigated the relative feature importance of lab test data, clinical data, and radiomics data.

### 3.7 Reliability quantification with conformal prediction

In the application of predicting ICU-admission of COVID-19 patients, besides the predictions themselves, the prediction reliability and uncertainty is of great importance in the management of health policies to optimize the usage of the limited ICU resources. Once the prediction reliability can be quantified, the ICU utilization policies can be made according to both how confident one has for a positive prediction and how severe the outcomes can be brought about by the current virus variant.

To quantify the prediction reliability, we leveraged the conformal prediction which was developed by Vladimir Vovk [42]. Conformal prediction assumes that the data abide by the independent and identical distribution (I.I.D) and outputs the credibility as the reliability information for each prediction. The applications and brief introduction of conformal predictors can be found in previous publications [36, 37, 43]. To compute the credibility, we leverage the conformal predictor based on the prediction probability and the steps can be summarized as the following steps:

1. Convert the predicted probability to a nonconformity measurement: a metric to quantify how well a particular feature-label combination conforms to the training data. Here, we leveraged a design of the nonconformity measurement *α*_*i*_ that has been validated in multiple machine learning applications [34, 44]:

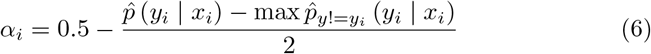 Here *y*_*i*_ and *x*_*i*_ denotes the label and feature of the *i − th* sample. In thisstudy, the predicted probability can be computed by SC, LGB or MLP.
2. Based on the nonconformity measurement, all the training samples’ non-conformity measurement values can be computed and the distribution will be further used to calibrate the credibility we have for a new prediction;
3. When making a new prediction, the nonconformity measurement *α*^∗^ for the test sample *x*^∗^ is computed based on the previous equation. Then, the P-value of the prediction, which indicates the credibility of the prediction is calculated by investigating the fraction of samples in the training distribution with larger nonconformity measurement:

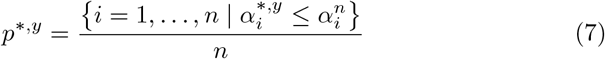 Here *p*^∗,*y*^ is the P-value of the assumed label *y* for the new sample *x*^∗^. It should be noted that, there are two P-values associated with the two labels (0: no ICU required, 1: need ICU) respectively.
4. The credibility of the prediction can be computed as the larger P-value [34], which reflects how well the most likely label conform to the distribution of the training data nonconformity measurement. If the credibility is low, the credibility we have for the prediction is low, which may provide us with a flexible tool to tune the policies in medical resource management.

In this study, to show the validity of the reliability quantification with conformal prediction and whether more reliable predictions mean higher likelihood of being correct, we used the conformal prediction with shrunken centroids (CPSC) algorithm and tested it on the HSS data set as an example (considering the stable performance and the lowest computational time of SC, and the relatively higher prevalence of positive cases in HSS data set) and performed two types of reliability analysis:

On the one hand, we partitioned the predictions into two categories based on the prediction credibility: unreliable predictions if the credibility is below a threshold *ϵ*, and reliable predictions if the credibility is above a threshold. Furthermore, within the reliable predictions, there are positive predictions and negative predictions. To make it simple to understand, the output of the conformal predictor is described as a tri-light system: red (reliable positive predictions), yellow (unreliable predictions) and green (reliable negative predictions). Then, we investigated the variations in AUROC, AUPRC, F1-score and the number of predictions within the red- and green-light predictions as the credibility threshold *ϵ* changes.

On the other hand, we investigated the credibility of the correct prediction, the false positive predictions and the false negative predictions and tested whether the correct predictions are assigned with higher credibility values.

For both analyses, to show the model performance robustness, we boot-strapped the training data for 30 times and reported the metrics within the 30 parallel experiments.

### 3.8 Ethics and registration

The protocol of this multi-center study was approved by the institutional review board of Jinling Hospital, Nanjing University School of Medicine (2020NZKY-005-02). The written informed consent was waived because this was a retrospective study and present no more than minimal risk of harm to subjects and involved no such procedures.

## 4 Results

### 4.1 Patient cohort

We collected 3,646 patients for analysis, including a training cohort (n = 1,451), a validation set (n = 662), an external test set (n = 1,263) based on the data collected from the Huoshenshan field hospital, and a specific test set (n = 270) based on Delta and Omicron variants 2. Prediction models were built for prediction of ICU admission (adverse cases in training set/validation set/HSS test set/Variants test set, n = 39/21/262/11, respectively). This cohort had 1,830 men (50.2%) and 1,816 women (50.8%), with a median age of 53.7 years (IQR, 42–65 years). The median age among men was 53.2 years (IQR, 41–65 years) and the median age among women was 52 years (IQR, 44–65 years). No statistical difference in age was found between men and women in this cohort.

### 4.2 Model prediction performance

The performance of the prediction models on the validation set, Huoshenshan (HSS) test set and the Delta and Omicron variants test set is shown in Table 1 and Figure 3. According to the results, the LGB model performs the best on the validation set while its performance can decrease sharply under data distribution drifts when the model is tested on the HSS field hospital data set and the Delta and Omicron variants data set. On the contrary, the SC model is the most robust model under data distribution drifts with the performance metrics more stable on the validation data set (AUROC: 0.78), HSS data set (AUROC: 0.77) and Variants data set (AUROC: 0.79).

**Table 1.** The performance of the ICU prediction model on three data sets over 30 bootstrapping experiments. Mean and 95% confidence interval are reported.

| Data set | Model | Feature Selection | Classifier Hyperparameter | AUROC | AUPRC |
| --- | --- | --- | --- | --- | --- |
| Validation | SC | LASSO $C = 0.01$ | $\Delta = 0.01$ | 0.803 [0.793, 0.813] | 0.221 [0.192, 0.250] |
| Validation | LGB | SC $\Delta = 0.05$ | $\alpha = 1, D = 9, N = 15$ | 0.861 [0.849, 0.873] | 0.306 [0.278, 0.334] |
| Validation | ANN | SC $\Delta = 0.4$ | layers=[50,30,20] | 0.755 [0.713, 0.800] | 0.156 [0.090, 0.256] |
| HSS | SC | LASSO $C = 0.01$ | $\Delta = 0.01$ | 0.743 [0.734, 0.753] | 0.462 [0.448, 0.477] |
| HSS | LGB | SC $\Delta = 0.05$ | $\alpha = 1, D = 9, N = 15$ | 0.705 [0.696, 0.715] | 0.438 [0.427, 0.449] |
| HSS | ANN | SC $\Delta = 0.4$ | layers=[50,30,20] | 0.731 [0.636, 0.788] | 0.431 [0.340, 0.513] |
| Variants | SC | LASSO $C = 0.01$ | $\Delta = 0.01$ | 0.746 [0.713, 0.779] | 0.172 [0.147, 0.197] |
| Variants | LGB | SC $\Delta = 0.05$ | $\alpha = 1, D = 9, N = 15$ | 0.774 [0.753, 0.794] | 0.131 [0.110, 0.153] |
| Variants | ANN | SC $\Delta = 0.4$ | layers=[50,30,20] | 0.768 [0.525, 0.874] | 0.219 [0.072, 0.423] |

**Fig. 3.**
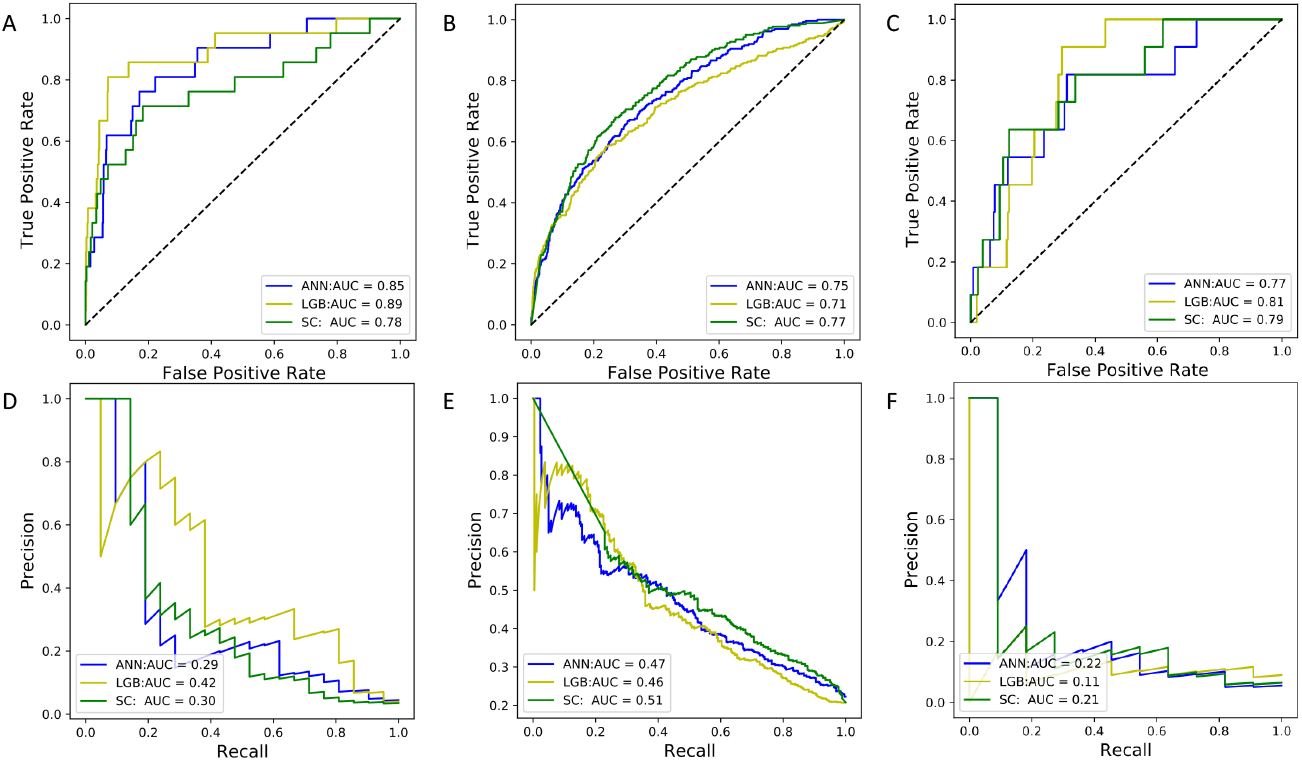
The receiver operating characteristic curves and precision recall curves of the models evaluated on the validation set (A,D), the Huoshenshan field hospital data set (B,E), and the Delta and Omicron variant data set (C,F).

### 4.3 Important feature analysis

To investigate the important features and their influence on the model decision, we leveraged the SC algorithm considering its high AUROC and AUPRC under data distribution drifts as well as its simplicity and interpretability based on the shrunken contrast introduced in Section 3D. After bootstrapping the training data for 30 times and taking a mean value of the shrunken contrast (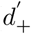 for the positive predictions and 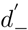 for the negative predictions) over the 30 parallel experiments, we report the feature importance of the three types of data: lab test data, clinical data and radiomic data in Fig. 4. The results indicate that the averaged feature importance of the lab test data is higher than that from the other two types of features (*p <* 0.01, Wilcoxon rank-sum test).

**Fig. 4.**
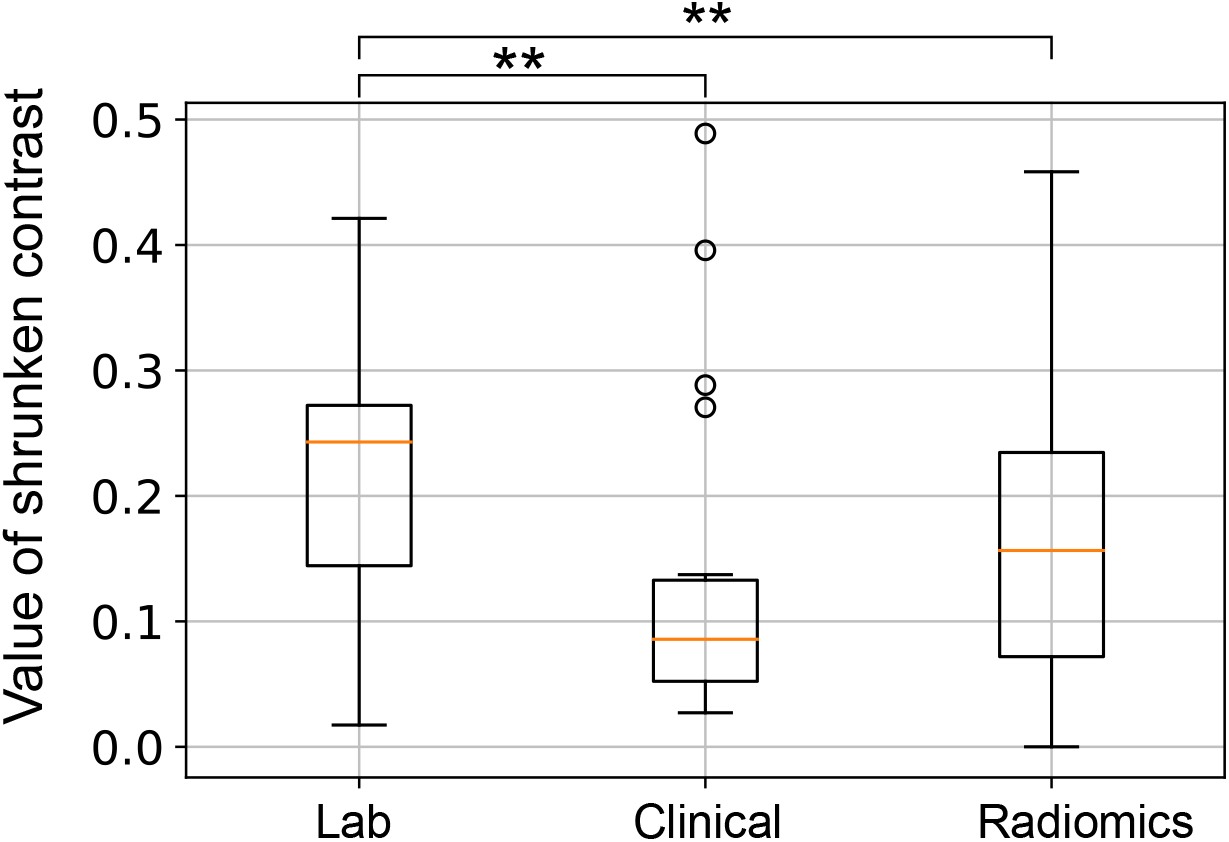
The feature importance quantified by the absolute values of the shrunken contrasts for the positive class centroid averaged over 30 times of bootstrapping experiments. The lab test data contains 19 features; the clinical data contains 18 features; the radiomic data contains 9913 features. Statistical significance: *: *p <* 0.05, **: *p <* 0.01, ***: *p <* 0.001.

Then, the most important features with an absolute value of shrunken contrast above 0.4 (d_+^−’” *>* 0.4) and the signs of the features associated with a positive prediction are reported in Table 2. Here a positive sign indicates that a higher value of a feature is more likely to lead to a positive prediction. The results show that the most important feature found by SC is the presence of dyspnea and the sign is positive, which indicates that the patients with dyspnea is more likely to be predicted as needing ICU admission. besides the clinical symptom of dyspnea and the lab test value of Lactate dehydrogenase, the majority of the important features are based on radiomics which suggests that CT radiomic features are the main features that are important in the model decision making process. Although the importance of the CT radiomics is relatively less evident than the lab test features when viewed as a group, due to the large number of radiomic features, there are still many features important in the model decision making process.

**Table 2.**
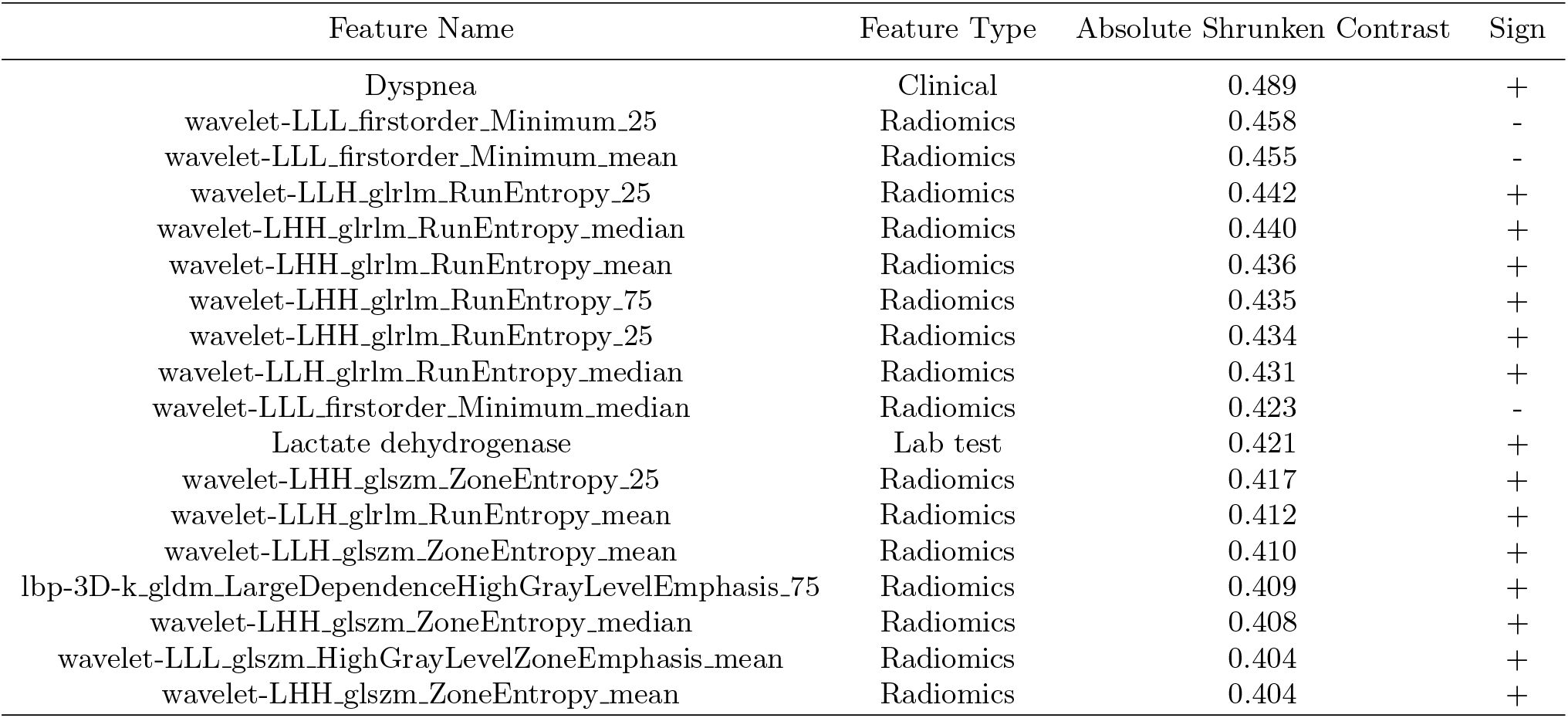
The most important features found by the shrunken centroids algorithms and their signs for making a positive prediction. The features were selected based on an absolute shrunken contrast above 0.4 averaged over 30 bootstrapping experiments. For the radiomic features, the last field indicates the statistics calculated over all lesions for a patient (e.g., 25: 25th percentile, 75: 75th percentile).

### 4.4 Prediction credibility analysis

To assist in the flexible medical resource management, besides giving the predictions, the conformal prediction framework was applied to enable the users to understand the reliability of each specific prediction made by the algorithms. The results of the two types of reliability analyses introduced in Section 2F are shown in Fig. 6. The results indicate that as the credibility threshold increases, the number of predictions decreases as the unreliable predictions with a credibility below the threshold are filtered out. Meanwhile, the model performance metrics: AUROC, AUPRC and F1-score increases accordingly when the more reliable predictions are investigated. The trend indicates that as the credibility requirement becomes stricter, the prediction performance is improved at the sacrifice of the number of predictions made by the algorithm, while *ϵ* = 0.3 can be a relatively balanced choice.

In addition, the mean credibility of the correct predictions, false positive predictions and false negative predictions are reported in Fig. 5. According to the results, the mean credibility of the correct predictions are significantly higher than those of the false positive predictions and false negative predictions (*p <* 0.001, Wilcoxon rank-sum test). To sum up, the two types of reliability analyses show that the credibility are positively related to the prediction performance: the higher the prediction credibility is, the more likely the prediction is correct, which enables the users to flexibly control the prediction performance via the prediction credibility.

**Fig. 5.**
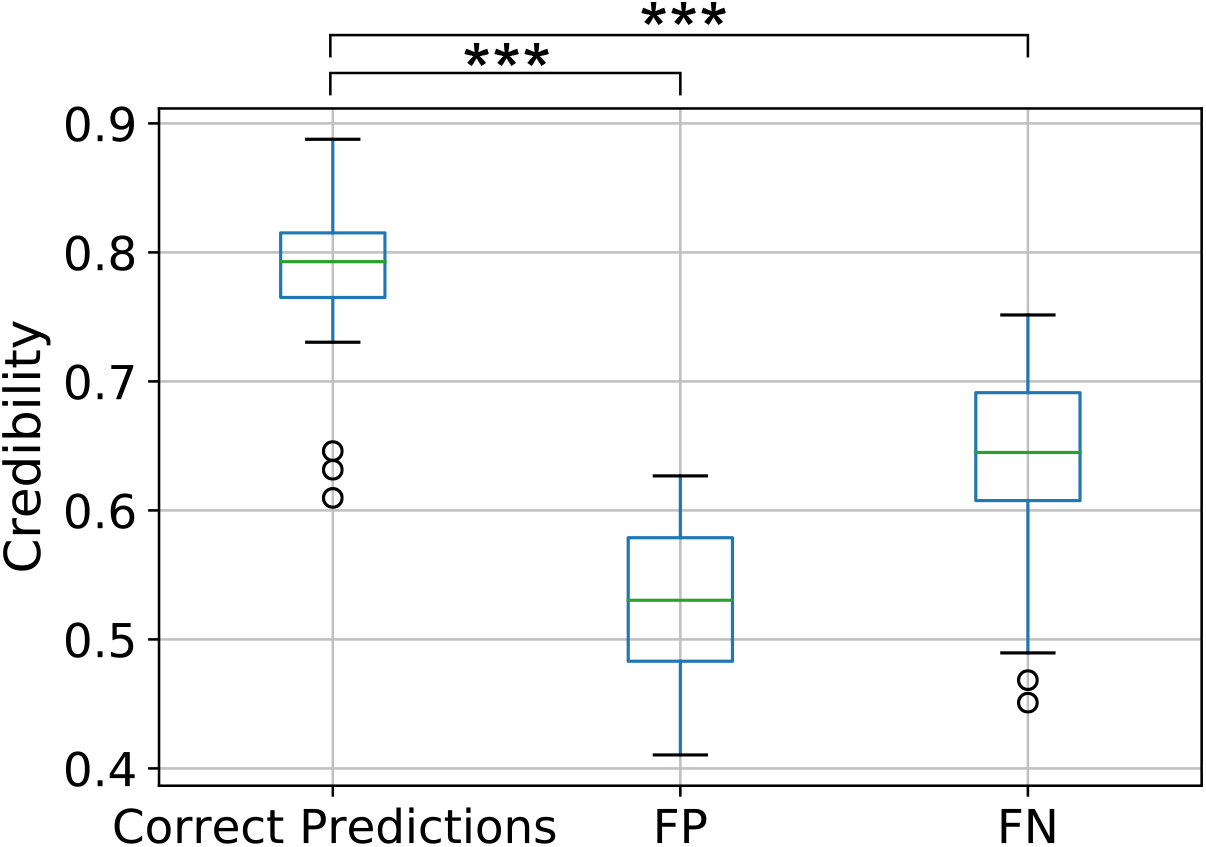
The mean credibility of different types of predictions. Correct predictions, false negative predictions (FN), and false positive predictions (FP) are reported. The mean credibility is calculated with data in the same prediction types in each bootstrapping experiment (30 times of bootstrapping experiments in total). Statistical significance: *: *p <* 0.05, **: *p <* 0.01, ***: *p <* 0.001.

**Fig. 6.**
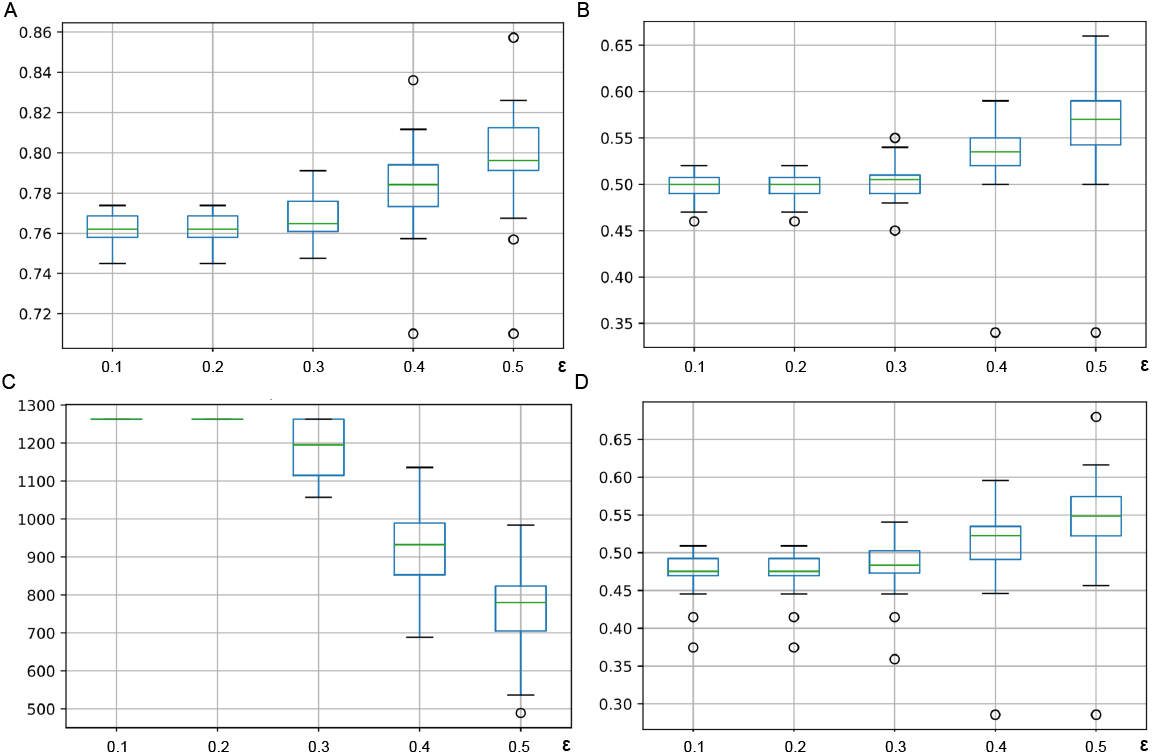
The model performance under varying threshold of credibility. The AUROC (A), AUPRC (B), number of predictions (C) made and F1-score (D) after the unreliable predictions are filtered out based on the varying credibility threshold *ϵ*. It should be noted 30 results from the bootstrapping experiments were reported.

## 5 Discussion and conclusion

Over the past two years, the number of COVID-19 patients has increased rapidly since outbreak of the pandemic, particularly with the spread of more contagious variants. This increase of COVID-19 patients has often overwhelmed the medical resources such as capacity problems in general wards and intensive care units (ICUs). To enable better medical resource allocation during this pandemic, the proposed model can predict whether a patient admitted to the general ward will be admitted to the ICU. Using multimodal data including CT radiomic features, clinical features and lab test features, these models were evaluated on the Huoshenshan field hospital data set, Delta and Omicron variant data, and the model performance was also tested under data distribution drifts. Furthermore, we have discovered key insights into the prediction reliability with the conformal prediction framework, enabling users to make a more informed decision regarding ICU resource allocation based on the prediction credibility.

Testing the performance of models predicting severity is especially of interest across mutational variants of COVID-19. Our results showed we were able to validate the ICU prediction models based on LGB, SC and ANN on the Huoshenshan field hospital test set and the Delta and Omicron variant test set, respectively. Among the ICU prediction models, the LGB model performed the best on the validation set while the simpler and more interpretable shrunken centroids (SC) model was generally the best on the two test data sets under data distribution drifts. The more flexible LGB and ANN models have high variance and overfit to the training data collected in the first wave of COVID-19 spread. Therefore, the stable performance of the SC model shows that this simpler model has the highest potential to also extrapolate to future variants.

To identify discriminating features for prediction, we found that the prescence of dyspnea, the lab test lactate dehydrogenase, three first order radiomic features, and 13 higher order (glrlm, glszm, gldm-based) radiomic features are important in the model decision making. Lactate dehydrogenase (LDH), a liver biochemistry marker related to liver impairment, and has been shown to be associated with poor prognosis for patients with COVID-19 [45][33]. The presence of dyspnea has been widely recognized as an indicator of the severity of COVID-19 and the potential admission to ICU. It has also been deemed by the Centers for Disease Control and Prevention as a severe symptom of COVID-19 as well as for any COVID vaccines. In our previous study [16] where an LGB model was used to analyze the important features in the prediction of ICU requirements, mechanical ventilation requirements and whether a patient would die from COVID-19 within 28 days, the majority of the important features are clinical features and lab test features including change of LDH and the presence of dyspnea [16]. However, in this study, the majority of important features found by the SC are radiomic features. We hypothesize that more radiomic features being important may be the main reason that enables the model to perform better than LGB under data distribution drifts. Considering the difference between Huoshenshan field hospitals and other hospitals in the handling of lab tests (e.g. within 3d after admission vs within 24h after admission) and the different clinical symptoms brought about by different COVID variants, the CT radiomic features are potentially more homogenous across different hospitals or for different COVID variants. Therefore, we suggest that CT radiomic features may be more reobust under data distribution drifts.

Conformal prediction enables the model to output not only the predicted labels but also the credibility of the predictions [42] [43]. We must clarify that the prediction reliability is not equally as the predicted probability. While predicted probability can be an intuitive indicator of a model’s confidence on a particular prediction [37], it relies heavily on model assumptions and does not give uncertainty information explicitly. By contrast, the credibility given by the conformal prediction better reflects prediction uncertainty because it relies on the statistical distribution of the nonconformity measurement in the training observations with the i.i.d. assumption that is less strict than the assumptions of most machine learning models. Changing the threshold on the prediction credibility is also relatively independent of changing the threshold on the predicted probability. The thresholding on the credibility can be used to quantify the confidence a user has for a prediction and whether or not to believe in the predictions made by the predictor, while the thresholding on predicted probability reflects what types of predictions are made by the model for a specific patient. To sum up, the prediction credibility and prediction probability can be regarded as two key types of prediction information. Based on the quantified credibility, a clinical center can flexibly adapt their health policies based on both the model predictions and the prediction credibility.

On top of the prediction credibility given by the conformal prediction framework, we also developed a “Red-Yellow-Green” tri-light warning system. For instance, the ICU resources can be allocated only to red-label patients[46], yellow-label patients need closer monitoring to determine the requirement of medical resources, while green-label patients are in a stable condition with a low probability of disease progression. Additionally, the tri-light system based on the prediction credibility can be adaptive to different virus variants.

It should be mentioned that, patients predicted yellow are not necessarily and in fact “safer” than the patients labeled red: the tri-light system can clearly inform the healthcare team that “the current medical information is not enough to make a reliable judgement for the prediction labeled yellow”. Therefore, more medical monitoring should be maintained. To emphasize, the dynamic setting of the tri-light system can leverage the conformal prediction framework to adaptively set health policies according to the different scenarios with the specific resources status and the forthcoming new variants in pandemic.

Although this study presents the design and development of a predictive model of ICU resource allocation for the COVID-19 patients, there are several limitations. Firstly, the current model is only designed to predict the requirement of ICU care rather than predicting specific needs of medical resources, such as mechanical ventilation or extracorporeal membrane oxygenation (ECMO) in severe patients. Although we have attempted to predict whether a patient admitted to the ICU will die based on the updated data upon the ICU admission (AUROC 0.77 and AUPRC 0.06 on HSS test set), the model performance was poor due to the potential reason of limited training data. In the future, as more ICU cases are collected across different medical institutions, more detailed patient outcome prediction models can be developed and the medical resource allocation after the ICU admission can be further optimized. Secondly, the effect of treatment was not considered in this study. Patients under appropriate treatment, including but not limited to the oxygen therapy, antiviral treatment and antibiotic treatment, can recover soon and the outcomes can be improved so that ICU admission may not be needed for a patient predicted to require ICU. Furthermore, although our study involves large sample sizes with clear prognosis information under different data distributions, the data were collected only from hospitals in China which can limit the generalizability of models into other areas considering the different protocols used across different countries.

In conclusion, we presented an end-to-end AI system based on conformal predictions to rapidly identify high-risk patients of COVID-19. The system outputs the prediction probability as well as the credibility of each prediction, so as to classify patients into high risk, low risk and uncertain risk. Importantly, we highlight that our method is applicable to both the original strain and more recent variants of COVID-19. Given the status of constant mutating of COVID-19, the continued growth of AI-driven techniques to identify epidemiological risks early will be key to improve prediction, prevention, and detection of future global health risks.

## Data Availability

The data that support the findings of this study are available on request from the corresponding author (G.M.L.). The data with participant privacy/consent are not publicly available due to hospital regulation restrictions.

https://github.com/LeoLee7/COVID_trilight

## 7 Code availability

The codes that support the findings of this study are available here: https://github.com/LeoLee7/COVID_trilight.

## 8 Acknowledgements

This work was supported by the Young Scientists Fund of the National Natural Science Foundation of China (grants No. 82202150 to Xiao Li).

## 9 Author information

These authors contributed equally: Chuan Junxu, Qinmei Xu, Liu Li.

### 9.2 Contributions

First draft was written by Chuan Junxu, Qinmei Xu, and Liu Li. Changsheng Zhou, Xiao Li, Rong Wang, Yanjun Wu, and Jiangtao Wang collected the data. Zhen Zhou and Zijian Xing contributed analysis tools. Chuan Junxu, Qinmei Xu, Liu Li, and Xianghao Zhan performed the analysis. Mu Zhou, Xianghao Zhan, Olivier Gevaert, and Guangming Lu Conceived the analysis and provided critical revisions.

### 9.3 Corresponding authors

Correspondence to Xianghao Zhan, Olivier Gevaert, and Guangming Lu.

